# Identification of expanded and interrupted *ATXN2* repeat expansions in Parkinson’s disease cohorts

**DOI:** 10.1101/2025.03.26.25324719

**Authors:** Longfei Wang, Michael Milton, Liam G. Fearnley, Oneil Bhalala, Melanie Bahlo, Haloom Rafehi

## Abstract

Repeat expansions (REs) may be Parkinson’s disease (PD) risk factors. We screened whole genome sequencing data from the AMP PD Lewy Body Dementia (LBD) and PD cohorts for 37 REs associated with other neurological disorders, and identified both interrupted and uninterrupted CAG REs in *ATXN2* in 4/2,431 PD and 2/2,468 LBD cases, but none in controls. These findings suggest pleiotropy for certain REs in PD.

## Main paper

Parkinson’s disease (PD) is a debilitating neurodegenerative disorder with a strong genetic component. Common genetic risk factors for sporadic PD have been identified at 78 independent genome-wide significant loci in a recent multi-ethnic genome-wide association study (GWAS)^1^.

Repeat expansions (REs) of short tandem repeats (STRs) are important genetic causes of many neurodegenerative disorders and are collectively referred to as repeat expansion disorders (REDs)^2,3^. While STR lengths are inherently variable between individuals, some STRs are pathogenic only if expanded above a locus-specific threshold. Longer REs are often associated with increased disease severity and reduced age of onset. This phenotypic variation in REDs can result in substantial underdiagnosis. One recent population study screened whole genome sequencing (WGS) data and identified an allele frequency for known RED alleles of 1 in 283 individuals, which is much higher than approximately 1 in 3000 individuals who have a RED diagnosis, suggesting underdiagnosis, incomplete penetrance, or both ^4^.

While no REs are currently identified as being specific causes of PD, parkinsonism is sometimes observed as a symptom of some REDs, such as Spinocerebellar ataxia 2 (SCA2), which is caused by REs greater than the pathogenic threshold of 33 CAG repeats in *ATXN2*, without any interruptions to the CAG sequence^3^. Disease pleiotropy has been previously described for REs in *ATXN2*, with intermediate expansions between 30-32 CAG repeats strongly enriched in ALS cohorts^5,6^. Notably, REs greater than 32 repeats in *ATXN2* in which the DNA sequence is interrupted with CAA repeats have been observed in multiple PD studies, although reports are conflicting ^7,8^. Other reports also implicate REs in *RFC1* in PD cases ^9,10^. One limitation of previous studies is the lack of understanding of the prevalence of such REs in the general population.

Genotyping STRs in short-read sequencing data is now both feasible and reliable for most of the known pathogenic STR loci, due to advances in bioinformatics methods which have been validated against current gold standard lab-based methods ^3,4^.

Our study into the contributions of RE in PD was performed on a large European cohort consisting of WGS from 7,835 individuals from the Accelerating Medicines Partnership program for Parkinson’s Disease (AMP PD). The combined PD cohorts consisted of 2,431 PD cases, 230 PD-like cases (including disorders such as multiple system atrophy and progressive supranuclear palsy) and 1,043 controls (Figure 1A). An additional Lewy Body Dementia (LBD) cohort from the AMP PD comprised 2,468 cases and 1,663 controls. We also compared estimated frequencies of genotypes determined by gnomAD for 7,487 European individuals, although not all loci are catalogued.

**Figure 1.**
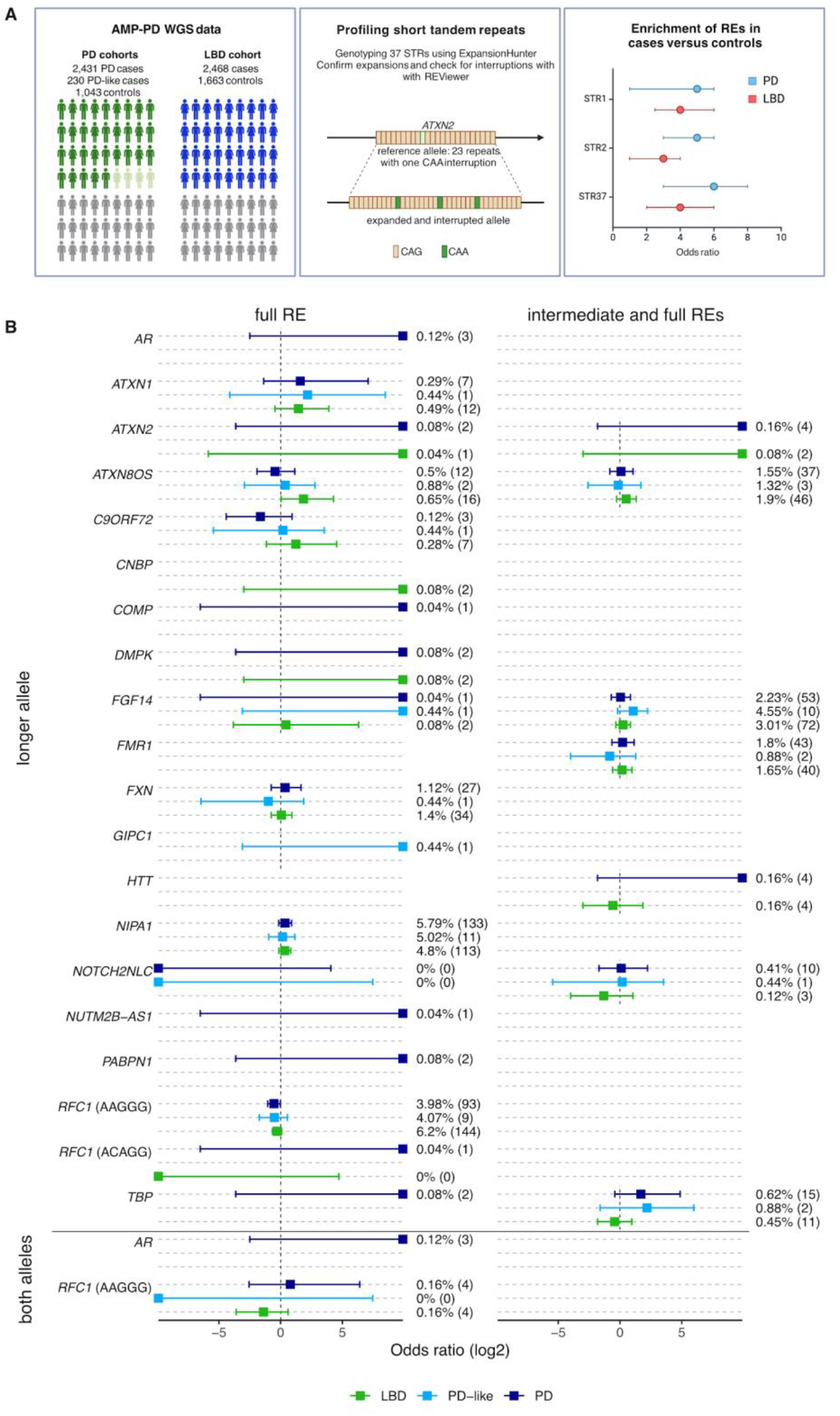
A) study design and overview. B) Bars indicate 95% confidence intervals. ORs to the far right are infinite ORs as there are 0 samples in the controls, while those on the far left are infinite ORs as the cases are 0. ORs for both alleles were only calculated for recessive REDs. Calculations for intermediate thresholds also include REs that are greater than the pathogenic threshold.

Genotyping was performed on a short-list of 37 STRs using ExpansionHunter ^11^. Alleles are referred to as REs if the genotype is larger than the reported number of repeats required to cause disease, i.e. the pathogenic threshold. A secondary, intermediate threshold was also applied to 10 loci based on previous reports that a shorter RE can also cause disease, typically with incomplete penetrance (Supplementary Table 1). Across both PD and LBD cohorts, we identified a combined 996 REs in the longer allele in the combined cases and controls in 19 genes (Supplementary Figure 1), with REs in *ATXN1, ATXN2, HTT, DM1, NIPA1* and *RFC1* being the most frequent, summarised in Supplementary Table 2. A further 464 REs in the intermediate range in six genes were also detected. We also observed 19 biallelic or hemizygous carriers of REs in *RFC1* (16) and *AR* (3 hemizygous males), both of which cause recessive REDs (Supplementary Figure 2). Using Fisher’s exact test, no statistically significant enrichment in cases versus controls was observed following false discovery rate correction (Figure 1B), indicating no extreme enrichment of REs in PD cases and acknowledging the modest sample size for detection of rare REs.

The CAG RE in *ATXN2* is the most widely reported STR associated with PD ^8,12^. We observe expansions with >32 repeats in 0.16% (4/2,431) of PD cases, 0.08% (2/2,468) of LBD cases, but with no expansions observed in either the PD (0/1,043) or LBD (0/1,663) control cohorts (Figure 2A). In gnomAD, 0.03% (3/9,487) of European individuals have intermediate *ATXN2* expansions between 33-34 repeats. *ATXN2* REs >32 were found in 32/59,564 (0.05%) individuals of European ancestry from 100,000 Genomes Project and TOPMed^4^. When considering only full REs > 34 repeats, the incidence is 0.08% (2/2,431) of PD cases, 0.04% (1/2,468) of LBD cases, and 0% n the PD and LBD controls, 0% in gnomAD and 10 in 59,564 (0.02%) in the combined 100,000 Genomes Project and TOPMed cohort^4^. Inspection of the *ATXN2* locus with REViewer identified that five of the six expanded alleles identified in the PD and LBD cohorts were interrupted with a CAG>CAA change. One PD case had 34 uninterrupted repeats, which is in the range for incompletely penetrant SCA2 (Table 1). In gnomAD, we identified two interrupted REs with 33 repeats and one uninterrupted RE with 34 repeats. While these findings do not reach statistical significance due to the small study sample size, they are consistent with previous reports that expanded and interrupted *ATXN2* REs are genetic contributors to PD risk. Characterisation of the longer allele across all individuals in the PD and LBD cohorts shows that the most common allele length is 22 repeats (Figure 2B). Next, we took a random stratified sample from the length distribution of the longer allele, examining 106 alleles to determine whether interruptions in the *ATXN2* STR are common. We show that all but two alleles had at least one CAG>CAA interruption (Figure 2C).

**Table 1.** Summary of REs in *ATXN2*. *carrier status as reported by AMP PD. NA - data not available, ^pathologically confirmed (moderate plaques (CERAD = C); extensive tangles (Braak NFT = 5), DNA obtained from brain tissue). HBS cohort scores are UPDRS, not MDS-UPDRS.

| ID | Patient 1 | Patient 2 | Patient 3 | Patient 4 | Patient 5 | Patient 6 |
| --- | --- | --- | --- | --- | --- | --- |
| Number of repeats | 39 | 37 | 37 | 34 | 34 | 33 |
| Number of interruptions<br>(CAG>CAA) | 4 | 4 | 5 | 0 | 1 | 1 |
| Cohort | HBS | LBD | PDBP | PDBP | PDBP | LBD |
| Diagnosis | PD | LBD^ | PD | PD | PD | LBD |
| Age range (y) | 50-59 | 60-69 | 60-69 | 70-79 | 60-69 | 70-79 |
| Family history of PD | Unknown | No | No | Yes | Yes | No |
| Carrier status* | No | No | GBA (N370S,<br>rs76763715),<br>APOE (E3/E4) | LRRK2<br>(G2019S,<br>rs34637584) | No | No |
| MDS-UPDRS I | 1 | NA | 3 | 6 | 19 | NA |
| MDS-UPDRS II | 6 | NA | 8 | 3 | 25 | NA |
| MDS-UPDRS III | 18 | NA | 11 | 24 | 31 | NA |
| MDS-UPDRS IV | 2 | NA | 0 | 4 | 0 | NA |
| Cognitive impairment | Nil<br>(MMSE = 30) | NA | Mild<br>(MoCA = 25) | Nil<br>(MoCA = 29) | Mild<br>(MoCA = 19) | NA |
| Olfactory impairment | NA | NA | Severe<br>(UPSIT = 11) | Nil<br>(UPSIT = 37) | Nil<br>(UPSIT = 35) | NA |

**Figure 2.**
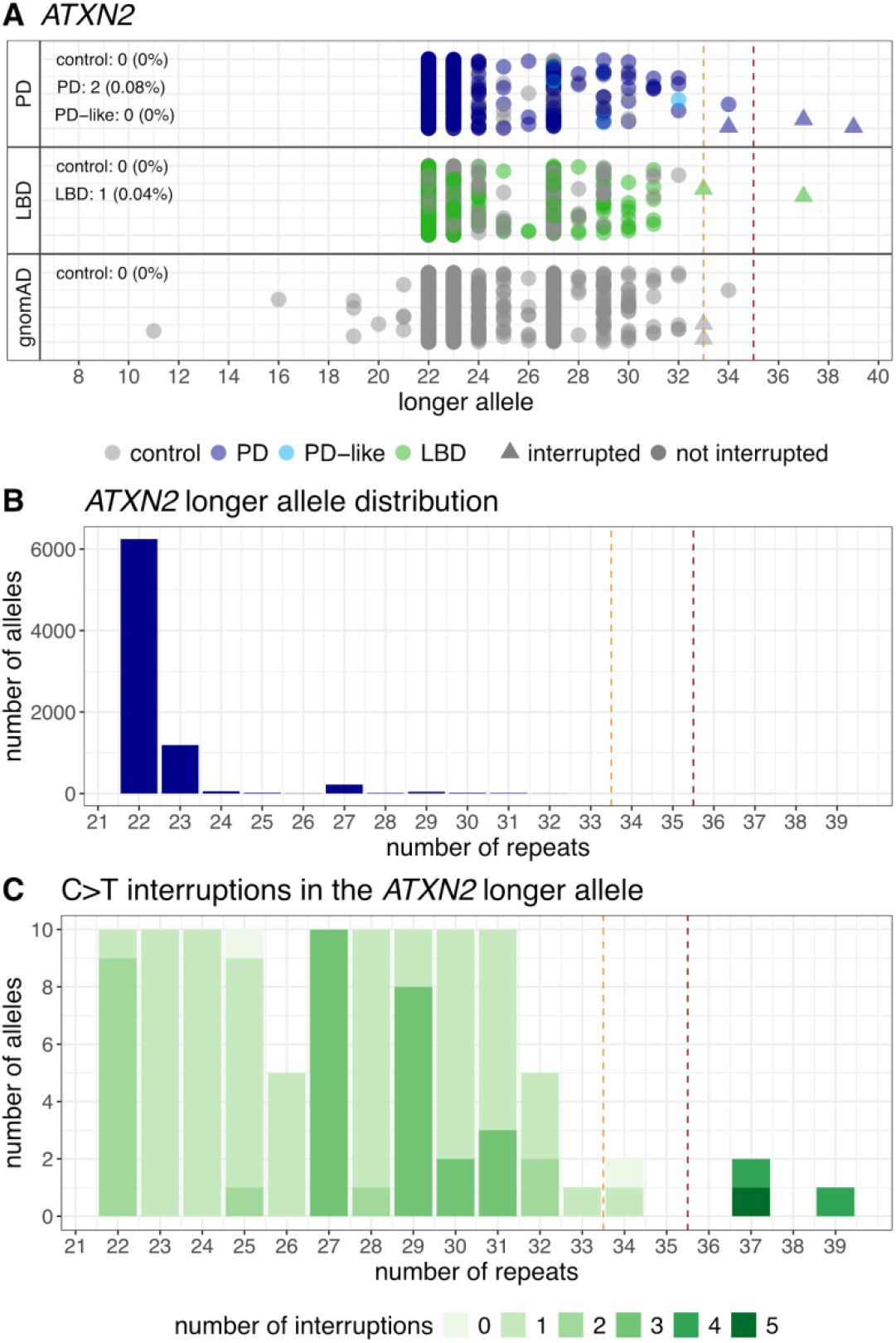
CAG REs detected in *ATXN2*. (A) Swimlane plot of the number of repeats in the longer allele in the PD, LBD and gnomAD cohorts. Triangles indicates REs with confirmed interruptions (only REs >= the intermediate threshold of 33 repeats were screened). Percentages indicate the number of individuals with a RE >= the pathogenic threshold of 35 repeats. (B) Distribution of the longer allele repeat size in all samples from PD and LBD cohorts. (C) Profiling CAG>CAA interruptions in a randomly generated and stratified subset of 106 individuals from the PD and LBD cohorts. Red dashed line: the pathogenic threshold at 35 repeats. Orange dashed line: pathogenic threshold at 33 repeats.

Recent studies have suggested a rare role for biallelic *RFC1*-AAGGG REs in PD, finding three biallelic carriers in a Finnish cohort of 569 individuals with PD (0.53%)^13^, an additional three cases in a follow up study in a Finnish cohort of 273 early-onset PD cases (1.1%) and four biallelic carriers in the Parkinson’s Progression Markers Initiative (PPMI) cohort looking at 903 (0.44%) non-Finnish Europeans with PD, but not in controls^14^. The PPMI is also a subset of the AMP PD and is included in the present study. Although biallelic individuals are observed in this study, with 0.16% (4/2,431) identified in the PD cohort, 0/230 in the PD-like cohort and 0.16% (4/2,468) in the LBD cohort, biallelic expansions were also present in control cohorts: 0.1% (1/1,043) in PD controls and 0.42% (7/1,663) in LBD controls, and 0.11% (8/7,487) in gnomAD. Therefore, no enrichment was observed. This is consistent with reports of 44 biallelic REs in *RFC1* in 29,496 (0.15%) European individuals in the 100,000 Genomes Project^4^. We note the discrepancy between the aforementioned PPMI study which identified four biallelic carriers in the PPMI - in our study, we only identified three cases from PPMI - the fourth case reported in the paper was determined to not be of European ancestry and was excluded in our analysis.

We observe 24 individuals with CAG REs >38 repeats in *ATXN1* in both cases and controls, with no significant enrichment (Figure 1). While uninterrupted CAG REs >38 repeats typically indicate SCA1, all REs observed contain at least 2 C>A interruptions, which negates the RE pathogenicity (Supplementary Table 1). Similarly, many REs were observed in *HTT* and *ATXN8OS* in both cases and controls, with no significant enrichment. Other REs, such as in the genes *GIPC1, AR, CNBP* and *DMPK* were observed in cases but not controls, however the incidence is very low and therefore no conclusions can be drawn.

To determine whether REs are enriched in PD and LBD overall, we performed Fisher’s Exact test on REs on three groups of data: 1) a merge of all loci, 2) all ataxia loci and 3) non-ataxia loci. No significant association was identified after False Discovery Rate (FDR) correction. In addition, we did not observe enrichment of REs in young-onset (<50 years) PD or LBD compared to late-onset, although again this analysis was limited due to small sample sizes, especially in the early onset groups. Meta-analysis was performed using Fisher’s method for combining p-values to determine whether there is a trend for RE enrichment across the PD and LBD cohorts, however no significant results were identified after FDR correction.

In conclusion, we identify REs in both PD and LBD cohorts, however there is no overall statistically significant RE enrichment in cases compared to controls. Many recent studies have shown that the prevalence of disease-causing REs is likely much higher than previously anticipated in general populations, which likely accounts for some of the associations of REs with PD observed in previous studies. Our finding of interrupted *ATXN2* REs in the PD cases but not controls follows the trend reported in the literature, although larger cohorts are required to achieve statistical significance for such rare variants. We also report for the first time interrupted *ATXN2* REs in LBD cases, suggesting a potential shared genetic mechanism.

## Methods

### Cohorts

The Walter and Eliza Hall Institute of Medical Research Human Research Ethics Committee (HREC 22/19) approved the study and all the procedures undertaken were in accordance with the ethical standards of the responsible committees.

This study was performed on the tier two data from the Accelerating Medicines Partnership program for Parkinson’s Disease (AMP PD, https://amp-pd.org/). Whole genome sequencing was performed by Macrogen and the Uniformed Services University of Health Sciences using the Illumina HiSeq XTen sequencer and was aligned to the GRCh38 reference genome. All primary analysis was performed within the Terra platform. For this study, we accessed WGS and phenotypic data from the LBD cohort and five PD cohorts: HBS, PDBP, PPMI, STEADY-PD3 and SURE-PD. Analysis was restricted to individuals of European ancestry due to population size constraints. Ancestry analysis was performed using reference populations from the 1000 Genomes Project ^15^ and a cohort of Ashkenazi Jewish individuals ^16^. The cohorts were merged with PLINK after excluding variants with a MAF < and LD pruning and excluding mismatching SNPs. In addition, 402 controls from the PD cohorts were reported to have family history of PD and were excluded from further analysis. Only unrelated individuals were included for further analysis. Relatedness analysis was performed with KING (v2.3.0). First, the merged pgen format genotype data was converted to bed format and pruned to exclude alleles with MAF<0.05 using plink2 (v2.00a3.7LM). Next, KING was run to identify individuals with up to fourth degree relatives. Related individuals were removed using a minimum weighted vertex cover method (MWVC). Priority was given to the samples in relatedness clusters based on their disease status in the following order: PD cases > PD-like cases > controls. As a result, 174 individuals were excluded from the PD cohorts (146 controls, 11 PD-like and 17 PD), and 71 from the LBD cohort (63 controls, 8 LBD cases).

In total, the study included 7,835 individuals of European ancestry, of which the combined PD cohorts consisted of 2,431(1,524 male, 907 female) PD cases, 230 (136 male, 94 female) PD-like cases (including disorders such as multiple system atrophy and progressive supranuclear palsy) and 1,043 (469 male, 574 female) controls, and the LBD cohort with 2,468 (1,555 males and 913 females) cases and 1,663 (841 males and 822 females) controls.

STR genotype distribution data were obtained from gnomAD from 7,487 non-Finnish European individuals as an additional control cohort. GnomAD contains genotype distributions for all loci included in this study, except for *FGF14, RILPL1, THAP11, ZFHX3* and *TAF1*.

### Repeat expansion genotyping

Repeat expansion genotyping was performed within the Terra platform using ExpansionHunter v5.0.0^11^. A short-list of 37 STRs (summarised in Supplementary Table 1) was curated on the following criteria: 1) the REDs are typically adult-onset disorders, 2) the main phenotype is a movement disorder or dementia. The final list consists of 31 autosomal dominant disorders, four autosomal recessive disorders, one X-linked dominant disorder and one X-linked recessive disorder. All alleles have a pathogenic threshold above which they are considered to be pathogenic REs. A secondary intermediate RE threshold was also applied to 10 loci based on reports that a shorter RE can also cause disease, typically with incomplete penetrance (Supplementary Table 1). For the *FGF14* locus, a lower pathogenic threshold was applied, as it has previously been shown that the genotype obtained from REs estimated from WGS data are highly accurate when repeats are <150 bp in length but are underestimated in size for larger REs with the discrepancy increasing exponentially with increasing RE size ^4,17^. REViewer was also used to confirm the motif for some loci, and to eliminate false positive RE calls ^18^.

Post-processing was performed on the ExpansionHunter calls to remove genotypes where the motif does not match the motif specified in the STR catalogue. This was performed using the package EH5_kmerfilter (https://github.com/bahlolab/EH5_kmerfilter). Briefly, this method used a kmer-based filtering to identify and exclude genotypes in which the dominant motif at a locus does not match the locus defined in the catalogue.

REs for a subset of loci (*ATXN1, ATXN2, ATXN8OS, C9ORF72, CNBP, DMPK, FMR1, HTT, RFC1, TBP*) which are known to have a high false positive rate or where interruptions are important above the pathogenic or intermediate threshold were visually checked for false positives or interruptions with REViewer plots. For example, *FMR1* is known to have a high false positive rate with ExpansionHunter, therefore all REs above the intermediate threshold were checked for false positives.

### Statistical analysis

Statistical analysis and graphical assessment was performed in R (v4.4.1). Fisher’s Exact test was used to determine potential enrichment of REs in cases versus controls for both the AMP PD and LBD cohorts. Enrichment was tested in the following ways in both the AMP PD and LBD cohorts:

1. In individual RE loci with dominant and recessive models and accounting for sex for X-linked loci,
  a. All cases versus controls
  b. All cases versus controls <50 years at baseline
  c. All cases versus controls >=50 years old at baseline
  d. In cases (PD, PD-like and LBD) only, comparing RE enrichment in early-onset (<50 years) versus late onset (>=50 years).
2. Group analysis in which loci are grouped into the following groups in cases versus controls:
  a. all loci,
  b. a grouped analysis of all ataxia RE loci and
  c. a grouped analysis of all non-ataxia RE loci.

Multiple test correction was performed using the False Discovery Rate method ^19^.

Meta-analysis was performed on the individual locus analysis with all samples for cases versus controls using Fisher’s method for combining p-values.

## Data availability

Access to the AMP PD data is available through the Terra platform (https://amp-pd.org/register-for-amp-pd). Individual-level genotype data, generated from the AMP PD, have been made accessible to the broader scientific community through the Terra workspace (https://app.terra.bio/#workspaces/bahlo_lab_amp_pd/MJFF-021399/files).

## Acknowledgements

This study was funded by the Michael J Fox Foundation for Parkinson’s Research (MJFF) and the Shake It Up Australia Foundation (MJFF-021399). The funder played no role in study design, data collection, analysis and interpretation of data, or the writing of this manuscript. H.R. was supported by an NHMRC Investigator Emerging Leadership 1 grant (1194364) and M.B. was supported by an NHMRC Investigator Leadership 1 grant (1195236). Additional funding was provided by the Independent Research Institute Infrastructure Support Scheme, the Victorian State Government Operational Infrastructure Program and the Murdoch Children’s Research Institute.

Data used in the preparation of this article were obtained from the Accelerating Medicine Partnership® (AMP®) Parkinson’s Disease (AMP PD) Knowledge Platform. For up-to-date information on the study, visit https://www.amp-pd.org.

The AMP® PD program is a public-private partnership managed by the Foundation for the National Institutes of Health and funded by the National Institute of Neurological Disorders and Stroke (NINDS) in partnership with the Aligning Science Across Parkinson’s (ASAP) initiative; Celgene Corporation, a subsidiary of Bristol-Myers Squibb Company; GlaxoSmithKline plc (GSK); The Michael J. Fox Foundation for Parkinson’s Research ; Pfizer Inc.; AbbVie Inc.; Sanofi US Services Inc.; and Verily Life Sciences.

ACCELERATING MEDICINES PARTNERSHIP and AMP are registered service marks of the U.S. Department of Health and Human Services.

Clinical data and biosamples used in preparation of this article were obtained from the (i) Michael J. Fox Foundation for Parkinson’s Research (MJFF) and National Institutes of Neurological Disorders and Stroke (NINDS) BioFIND study, (ii) Harvard Biomarkers Study (HBS) and the Stephen & Denise Adams Center for Parkinson’s Disease Research of Yale School of Medicine (CPDR-Y), (iii) National Institute on Aging (NIA) International Lewy Body Dementia Genetics Consortium Genome Sequencing in Lewy Body Dementia Case-control Cohort (LBD), (iv) NINDS Parkinson’s Disease Biomarkers Program (PDBP), (v) MJFF Parkinson’s Progression Markers Initiative (PPMI), and (vi) NINDS Study of Isradipine as a Disease-modifying Agent in Subjects With Early Parkinson Disease, Phase 3 (STEADY-PD3) and (vii) the NINDS Study of Urate Elevation in Parkinson’s Disease, Phase 3 (SURE-PD3).

BioFIND is sponsored by The Michael J. Fox Foundation for Parkinson’s Research (MJFF) with support from the National Institute for Neurological Disorders and Stroke (NINDS). The BioFIND Investigators have not participated in reviewing the data analysis or content of the manuscript. For up-to-date information on the study, visit michaeljfox.org/news/biofind.

Genome sequence data for the Lewy body dementia case-control cohort were generated at the Intramural Research Program of the U.S. National Institutes of Health. The study was supported in part by the National Institute on Aging (program #: 1ZIAAG000935) and the National Institute of Neurological Disorders and Stroke (program #: 1ZIANS003154).

The Harvard Biomarker Study (HBS) is a collaboration of HBS investigators [full list of HBS investigators found at https://www.bwhparkinsoncenter.org/biobank/ and funded through philanthropy and NIH and Non-NIH funding sources. The Stephen & Denise Adams Center for Parkinson’s Disease Research of Yale School of Medicine is funded through philanthropy and NIH and non-NIH funding sources. The HBS and CPDR-Y Investigators have not participated in reviewing the data analysis or content of the manuscript.

PPMI is sponsored by The Michael J. Fox Foundation for Parkinson’s Research and supported by a consortium of scientific partners: [list the full names of all the PPMI funding partners found at https://www.ppmi-info.org/about-ppmi/who-we-are/study-sponsors]. The PPMI investigators have not participated in reviewing the data analysis or content of the manuscript. For up-to-date information on the study, visit www.ppmi-info.org.

The Parkinson’s Disease Biomarker Program (PDBP) consortium is supported by the National Institute of Neurological Disorders and Stroke (NINDS) at the National Institutes of Health. A full list of PDBP investigators can be found at https://pdbp.ninds.nih.gov/policy. The PDBP investigators have not participated in reviewing the data analysis or content of the manuscript.

The Study of Isradipine as a Disease-modifying Agent in Subjects With Early Parkinson Disease, Phase 3 (STEADY-PD3) is funded by the National Institute of Neurological Disorders and Stroke (NINDS) at the National Institutes of Health with support from The Michael J. Fox Foundation and the Parkinson Study Group. For additional study information, visit https://clinicaltrials.gov/ct2/show/study/NCT02168842. The STEADY-PD3 investigators have not participated in reviewing the data analysis or content of the manuscript.

The Study of Urate Elevation in Parkinson’s Disease, Phase 3 (SURE-PD3) is funded by the National Institute of Neurological Disorders and Stroke (NINDS) at the National Institutes of Health with support from The Michael J. Fox Foundation and the Parkinson Study Group.

For additional study information, visit https://clinicaltrials.gov/ct2/show/NCT02642393. The SURE-PD3 investigators have not participated in reviewing the data analysis or content of the manuscript.

## Author contributions

H.R. and M.B. conceived and designed the research. H.R. and L.W. wrote the manuscript. M.M. and L.G.F. set up the pipelines for cloud computing (Terra). L.W. and H.R. performed the analysis, including individual-level analysis on cloud computing platforms (Terra). O.B. provided clinical interpretations of the results. M.B. provided funding, and resources. All authors read and approved the final manuscript.

