## Supplemental Figures for "Identification of expanded and interrupted *ATXN2* repeat expansions in Parkinson’s disease cohorts"

Supplementary materials

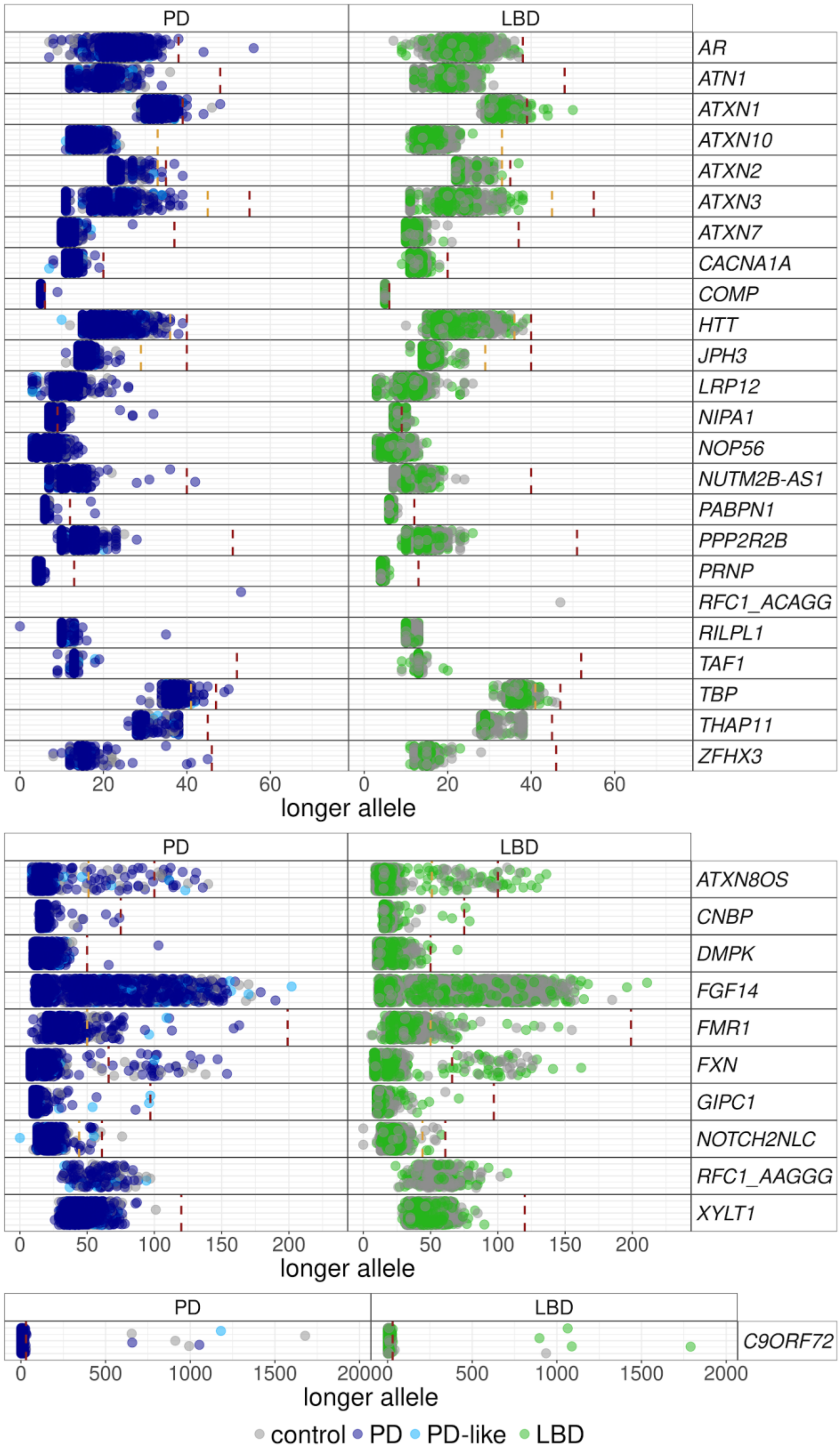


**Supplementary Figure 1. Swimlane plots showing the longer allele repeat numbers for the PD (blue), PD-like (light blue) and LBD (green) cases and controls (grey). Red dashed line: the pathogenic threshold. Orange dashed line: intermediate threshold.**

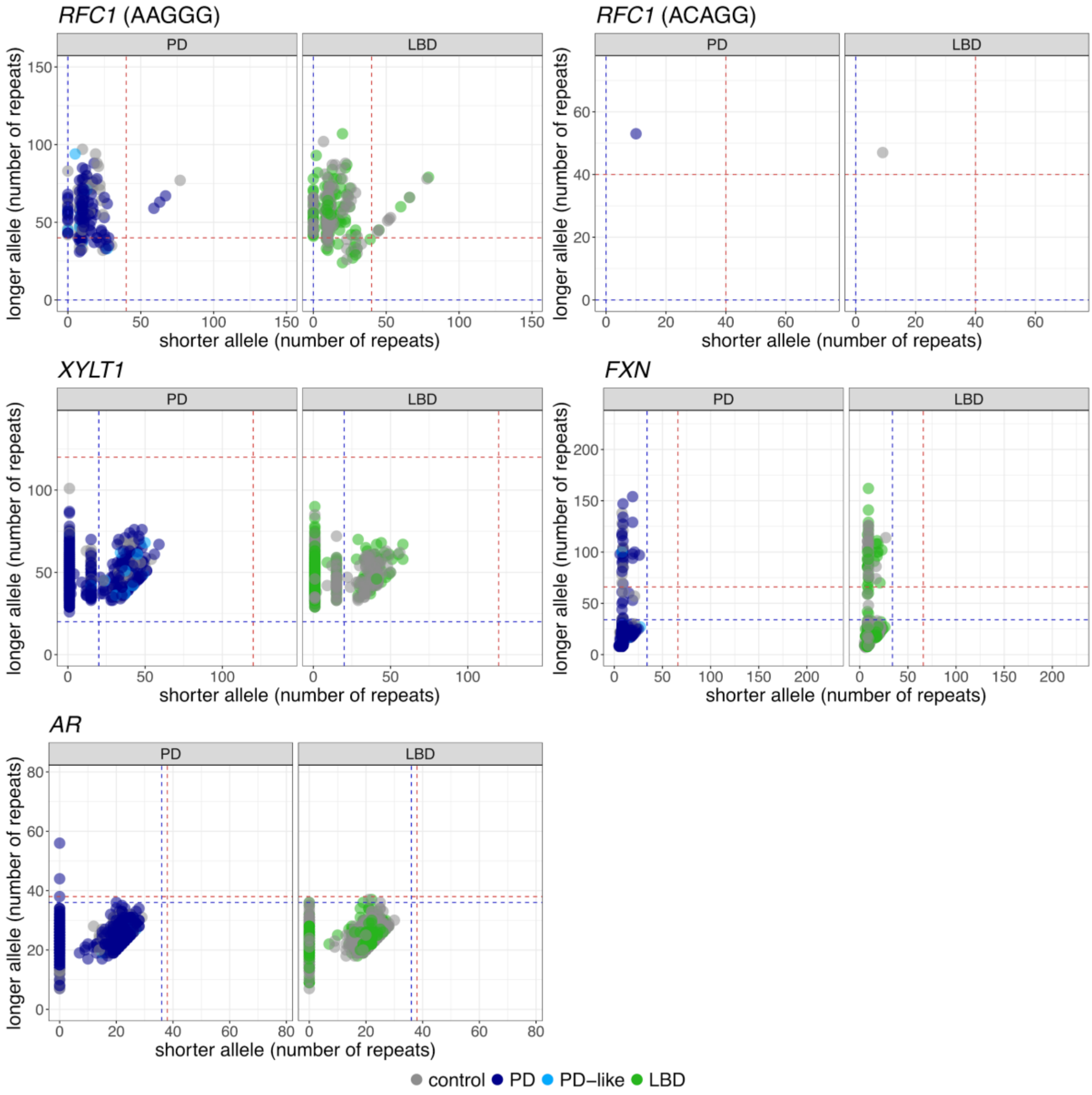


**Supplementary Figure 2.** Shorter and longer allele sizes estimated from EH5 for all recessively inherited disorders.
